# Predisposing factors associated with the severity of the illness in adults with Covid-19 in Nepal

**DOI:** 10.1101/2021.05.16.21257280

**Authors:** Roshan Kumar Jha, Anil Shrestha, Basant Tamang, Indu K.C., Shiv Kumar Sah

**Author notes:** **Corresponding author** Shiv Kumar Sah.

## Abstract

**Objective:** We aimed to determine the prevalence of the severity of COVID-19 illness and its associated predisposing factors in Nepal.

**Design:** Cross-sectional, observational study

**Setting:** Single-centered hospital-based study, conducted at Nepal armed police force (APF) hospital, Kathmandu, Nepal.

**Participants:** All individuals aged ≥18 years with laboratory-confirmed SARS-Cov-2 (the SARS-CoV-2 specific real-time-RT-PCR result positive), regardless the severity of their disease.

**Measurements:** Disease severity was evaluated as a primary outcome and age, sex, BMI, smoking history, alcohol history, Hypertension, diabetes mellitus were evaluated as predictors in the analysis.

**Results:** Mean ages of the patients were 40.79±16.04 years, and about two-thirds of the patients were male 146 (73.7%). More than half 57.1% (95%CI: 52.42-61.51) of the population had a mild infection, whereas 16.7% (95%CI: 7.4-24.6%) had severe/critical illness. In univariate analysis, each 1-year increase in age (OR: 1.05; 95% CI:1.030-1.081; P<0.001), each 1 unit increase in BMI (OR:1.12; 95% CI:1.02-1.25; P=0.033), comorbid illness (OR: 5.79; 95%CI: 2.51-13.33; P<0.001), hypertension (OR:5.95; 95%CI:2.66-13.30: P<0.001), diabetes mellitus (OR:3.26; 95%CI:1.30-8.15: P<0.005), and fever (OR:34.64; 95% CI:7.98-150.38; P<0.001) were independently associated with severity of the disease, whereas age (OR: 1.049; 95% CI: 1.019-1.080; P=0.02), hypertension (OR: 4.77; 95%CI: 1.62-14.04; P=0.004), and fever (OR: 51.02; 95%CI: 9.56-272.51; P<0.001) remained a significant predictive factors in multivariate analysis.

**Conclusion:** The majority of the patients with COVID-19 had a mild illness, with 16.7% severe illness. Age, BMI, hypertension, diabetes mellitus, comorbidity, and temperature were associated the severity of the illness. Age, hypertension, and fever emerged as an independent predictive factors in multivariate analysis, and thus, these vulnerable groups should be given special protection to the infection and proactive intervention should be initiated at an early stage of the infection to diminish the severity of the illness and improve the clinical outcome of the disease.

**Strengths and limitations of the study:**

- Much of the studies on COVID-19 in Nepal focus on the describing epidemiology and clinical profile of the disease, however, risk factors that contribute to the severity of the illness are overlooked.
- This study may help estimate the burden of the disease and identify the vulnerable group with poor prognosis, which is vital for clinicians and the public health approach to deal with the disease.
- Although limiting the study to a single-center with a relatively small sample size, it, however, allows evaluation of the importance of the demographic and geographical variation.
- Socio-economic factors, lifestyle, and availability of quality medical care may have contributed to the severity of the COVID-19, which needs to be addressed in a further large-scale study.

## Introduction

COVID-19, a new highly contagious ongoing outbreak of pneumonia, reported in Wuhan, Hubei province, China in late December 2019 (1-3), is associated with a novel beta-coronavirus, called severe acute respiratory syndrome coronavirus (SARS-CoV-2). COVID-19 has currently become a global health hazard to human society as the disease has strained the healthcare systems worldwide (4-7). The virus is highly transmissible from human to human (8), and the number of infected people increases at an exponential rate (9). The estimated case fatality rate ranges up to 17 % (10-13), and varies between the countries and across ages. WHO declared COVID-19 as a global pandemic on March 11, 2020 (14), and in Nepal, as of 26^th^ February 2021, over 2.5 million cases across the country have been detected, according to the ministry of health and Population (15).

COVID-19 is broadly classified into a range of clinical severity including, asymptomatic, mild, moderate (non-severe pneumonia), severe (severe pneumonia), and critical illness (16, 17). Epidemiological studies have indicated that of all individuals who become infected with SARS-CoV-2, a majority tend to have mild or no symptoms; however, an important minority will develop a predominantly respiratory disease that can lead to critical illness and death (1, 18, 19). A more recent data from a large cohort from China illustrated that 81% of patients had a mild disease while 14% severe and 5% developed critical illness (20).

The evidence on covid-19 and its causal agent SARS-CoV-2 has been rapidly accumulating, and recent studies across the regions have demonstrated that certain socio-demographic and clinical profile of the individuals may contribute to the severity of COVID-19, such as older age, male sex, and pre-existing hypertension, pulmonary disease, or cardiovascular disease (21-24). Importantly, because of the patient characteristics and the strain variation, these relative contributions of each trait to the risk for developing a more severe presentation of the COVID-19 might vary in different population groups or settings. In Nepal, to date, most clinical studies on COVID-19 have focused on describing the general epidemiologic and clinical characteristics. However, data on how the risks associated with demographic and underlying co-morbidities in this setting is lacking. Therefore the present study aims to determine the prevalence of severe disease and further underline the predictive factors associated with it.

## Methods

### Study design and setting

This was a cross-sectional observational study conducted at Nepal armed police force (APF) hospital, Kathmandu, Nepal, between December 2020 to January 2021. It is one of the most renowned hospitals in the country, containing 170 beds in Kathmandu valley, and amidst a dramatic surge in COVID-19 infection, it has become a leading hospital, serving general people with COVID-19 infections from all parts of the country.

### Study population and selection

All individuals aged ≥18 years with laboratory-confirmed SARS-Cov-2 (the SARS-CoV-2 specific real-time-RT-PCR result positive), regardless the severity of their disease were recruited for the study. Patients concurrently diagnosed with dengue fever, influenza A virus, Influenza B virus was excluded from the study.

### Clinical criteria for diagnosis of the disease and severity

COVID-19 was diagnosed according to the WHO interim guidance (25) who were admitted to the hospital regardless of the severity of the disease. A nasopharyngeal and throat swab samples were taken to test for SARS-CoV-2 by real-time reverse-transcription–polymerase-chain-reaction (RT-PCR) assay until two consecutive negative results at least 24 hours apart were achieved.

The severity of illness of each was assessed during hospitalization and classified into four clinical subtypes (i.e., mild, moderate, severe, or critically ill) based on management guidelines (26).

### Data collection procedure

Socio-Demographic, epidemiology, relevant clinical information, hospital courses, laboratory investigation, and treatment data of all consecutive laboratory-confirmed cases were extracted from medical records reviewed by attending physicians responsible for the patients with COVID-19 at Nepal armed police force Hospital, Nepal.

Collected information included demographic data (gender, age), signs and symptoms (fever, maximum temperature, cough, shortness of breath, fatigue, anorexia, muscle aches, headache, chills, nausea and vomiting, diarrhea, and confusion, etc.), pre-existing comorbid illness (hypertension, heart disease, diabetes, chronic obstructive pulmonary disease, liver disease, kidney disease, cancer etc), smoking and alcohol consumption history.

### Data processing and analysis

The data collected from patients’ medical records were entered into an excel spreadsheet and then exported to SPSS version 20 for analysis. Continuous variables were presented as mean□±□SD or median□±□interquartile range for skewed data. Categorical variables are presented as numbers and percentages in each category. Chi-square tests and Fisher’s exact (if cell count < 5) were performed to determine the significance of differences among categorical variables and differences between severe group and non-severe group using two-sample t-test or the Mann-Whitney U test depending on parametric or nonparametric data for continuous variables. Univariate and multivariate logistic regression models were used to explore the risk factors for severe cases. A p-value of <0.2 on univariate analysis were entered into a stepwise forward conditional multivariate logistic regression for the assessment of predictive factors associated with the severity of the disease. Statistical tests were two-tailed, and P<0.05 was considered significant.

### Ethical approval

The study was conducted in accordance with the Declaration of Helsinki. The study protocol was approved by Ethical Review Board (ERB) of Nepal Health Research Council (NHRC), Nepal (Ref. No: 1297). Informed consent was granted from all patients recruited in the study. Data were used anonymously to secure patients confidential integrity.

## Results

Table 1 demonstrates the demographic and clinical characteristics of the patients. The median ages of the patients were 40.79 (29.75-51.25) years, and a majority of the patients were male 146 (73.7%). Covid-19 cases were predominantly seen in Individuals having blood group O+ (34.34%) and A+ (33.33%). Seventy-six (76.4%) subjects had at least one comorbid illness, with predominantly hypertension 42 (21.2%), followed by diabetes mellitus 26 (13.1%), and 21 (10.6%) patients reported to have both hypertension and diabetes mellitus.

**Table 1.** Demographic and clinical characteristics of the patients

|  |  |
| --- | --- |
| n | 198 |
| Age ( IQR), years | 40.79 (29.75-51.25) |
| **Age distribution (years),n (%)** |  |
| 18-39 | 110 (55.6) |
| 40-59 | 59 (29.8) |
| ≥60 | 29 (14.6) |
| **Sex, n(%)** |  |
| Male | 146 (73.7) |
| Female | 52 (26.3) |
| BMI (IQR),kg/m<sup>2</sup> | 24.60 (22.0-27.60) |
| **Geography** |  |
| Himalayan region | 4 (2.00) |
| Hilly region | 177 (89.4) |
| Terai region | 17 (8.6) |
| **Education** |  |
| Secondary level (10-year education) | 95 |
| Higher secondary level (12-year education) | 44 |
| Bachelor degree | 27 (13.6) |
| Master's or above | 17(8.6) |
| Read or write | 15 (7.6) |
| **Profession** |  |
| Healthcare professionals | 63 (31.81) |
| Non-healthcare professionals | 135 (68.18) |
| **Blood group** |  |
| A+ | 66 (33.33) |
| B+ | 46 (23.23) |
| O+ | 68 (34.34) |
| AB+ | 16 (8.08) |
| O- | 2 (1.01) |
| **Type of infection** |  |
| Imported | 66 (33.3) |
| Local transmission | 132 (66.7) |
| **Contact history** |  |
| With suspected confirm | 192 (97.0) |
| Expose to similar illness | 4 (2.0) |
| Attend festival or mass gathering | 2 (1.0) |
| **Smoking history** |  |
| Smoker | 23 (11.6) |
| Non-smoker | 150 (75.8)) |
| Ex-smoker | 25 (12.6) |
| **Alcohol consumption** |  |
| Alcoholics | 35 (17.7) |
| Non -alcoholic | 137 (69.2) |
| Ex-alcoholic | 26 (13.1) |
| **Food habit, n(%)** |  |
| Vegetarian | 13 (6.6) |
| Non-vegetarian | 185 (93.4) |
| **Mask use,n(%)** |  |
| Yes | 183 (92.4) |
| No | 15 (7.6) |
| **Mask use types (n=183) ,n(%)** |  |
| Medical/ surgical | 122 (66.66) |
| Respirator | 7 (3.82) |
| Cloth mask | 54 (29.50) |
| **Comorbid illness (%)** | 76 (76.4) |
| Hypertension | 42 (21.2) |
| Diabetes mellitus | 26 (13.1) |
| CVD | 3 (1.5) |
| CKD | 1 (0.5) |
| Cancer | 2 (1.0) |
| Hypothyroidism | 3 (1.5) |
| COPD | 6 (3.0) |
| HTN+DM | 21 (10.6) |
| **Symptoms ,n (%)** |  |
| Present | 123 (62.1) |
| Absent | 75 (37.9) |
| Respiratory symptoms | 98(49.5) |
| Digestive symptoms | 14 (7.1) |
| Respiratory + digestive symptoms | 10(5.1) |
| fever | 82 (41.4) |
| Myalgia | 31 (15.7) |
| Headache | 39 (19.7) |
| Fatigue | 5 (2.5) |
| Sign | 79 (39.9) |
| Taste/smell | 47(23.7) |
| Throat congestion | 55 (27.8) |
| Tonsil swelling | 2 (1.0) |
| Unconsciousness | 1 (0.5) |
| Viral shedding time after the onset of illness, median(IQR), days | 8 (6-11) |
| Hospital days stay ,median (IQR), days | 11 (7-14) |

About two-thirds (62.1%) of patients were symptomatic, with about half (49.5%) of the subjects had respiratory symptoms, followed by digestive symptoms (7.1%). Among symptomatic individuals, 82 (41.4) subjects had a fever, followed by headache 39 (19.7%) and myalgia 31 (15.7%). Seventy-nine (39.9%) individuals presented with clinical signs, with more than half, 55 (27.8) reported having throat congestion, followed by taste/smell loss 47(23.7%). Median viral shedding time was 8 (6-11) days. Median hospital stay was 11 (7-14) days.

Table 2 demonstrates the prevalence of severity of the illness. More than half 57.1% (95%CI: 52.42-61.51) population had mild infection, while about one fourth 26.3% (95%CI: 19.2-32.85) had moderate, and 16.7% (95%CI: 7.4-24.6%) had severe/critical illness.

**Table 2.** prevalence of COVID-19 illness severity

|  | N =198 | Prevalence | 95%CI |  |
| --- | --- | --- | --- | --- |
|  |  |  | LL | UL |
| Mild | 113 | 57.1 | 52.42% | 61.51 |
| Moderate | 52 | 26.3 | 19.2 | 32.85 |
| Severe/critical | 33 | 16.7 | 7.4 | 24.6 |

Figure 1 illustrates the association between gender and the symptoms present in COVID-19 patients. Symptoms present in male subjects were significantly higher 98 (67.12%) than that seen in female counterparts 25(40.32%) (P=0.015)

**Figure 1.**
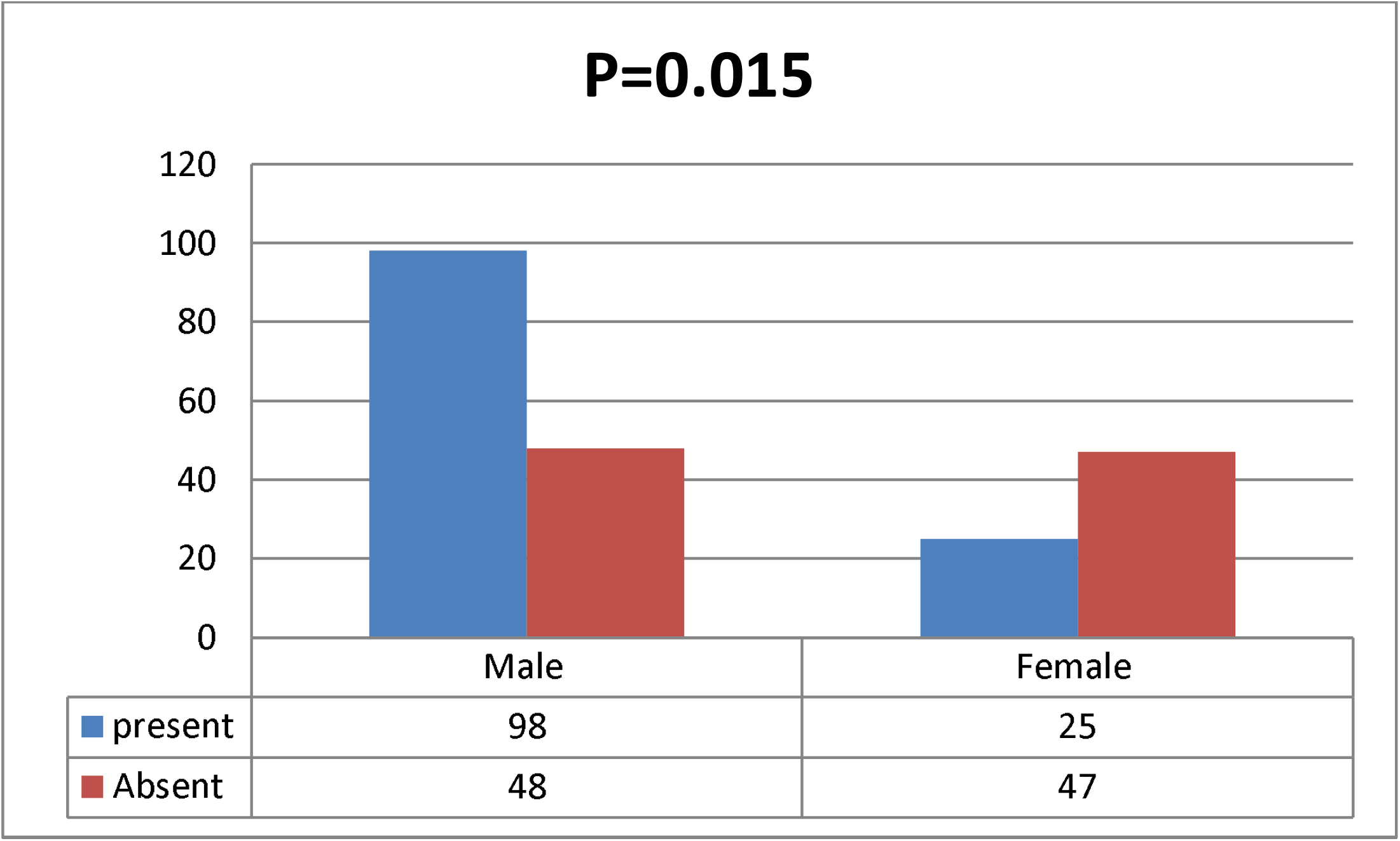
proportion of symptomatic cases by gender.

Table 3 illustrates the association of severity of the disease with socio-demographic and clinical parameters of the patients. Mean ages of the enrolled patients were significantly higher in patients with severe illness (53.21±16.99 years) compared to their mild (34.77±11.10 years) and moderate (46.86±17.35) counterparts (P<0.001). The severity of the disease was significantly linked to hypertension (P<0.001), and diabetes mellitus (P<0.001); in hypertension, the Proportion of covid-19 severe cases (54.5%) were predominantly higher compared to their mild (1.8%)and moderate (42.3%) counter parts, and in the diabetes mellitus, the Proportion of the severe cases were significantly higher (30.3%) compared to their mild (1.8) and moderate counterparts (26.9%). Viral shedding time and hospital stay days were higher in severe cases compared to mild and moderate cases; however, the difference was not significant (P>0.05).

**Table 3.**
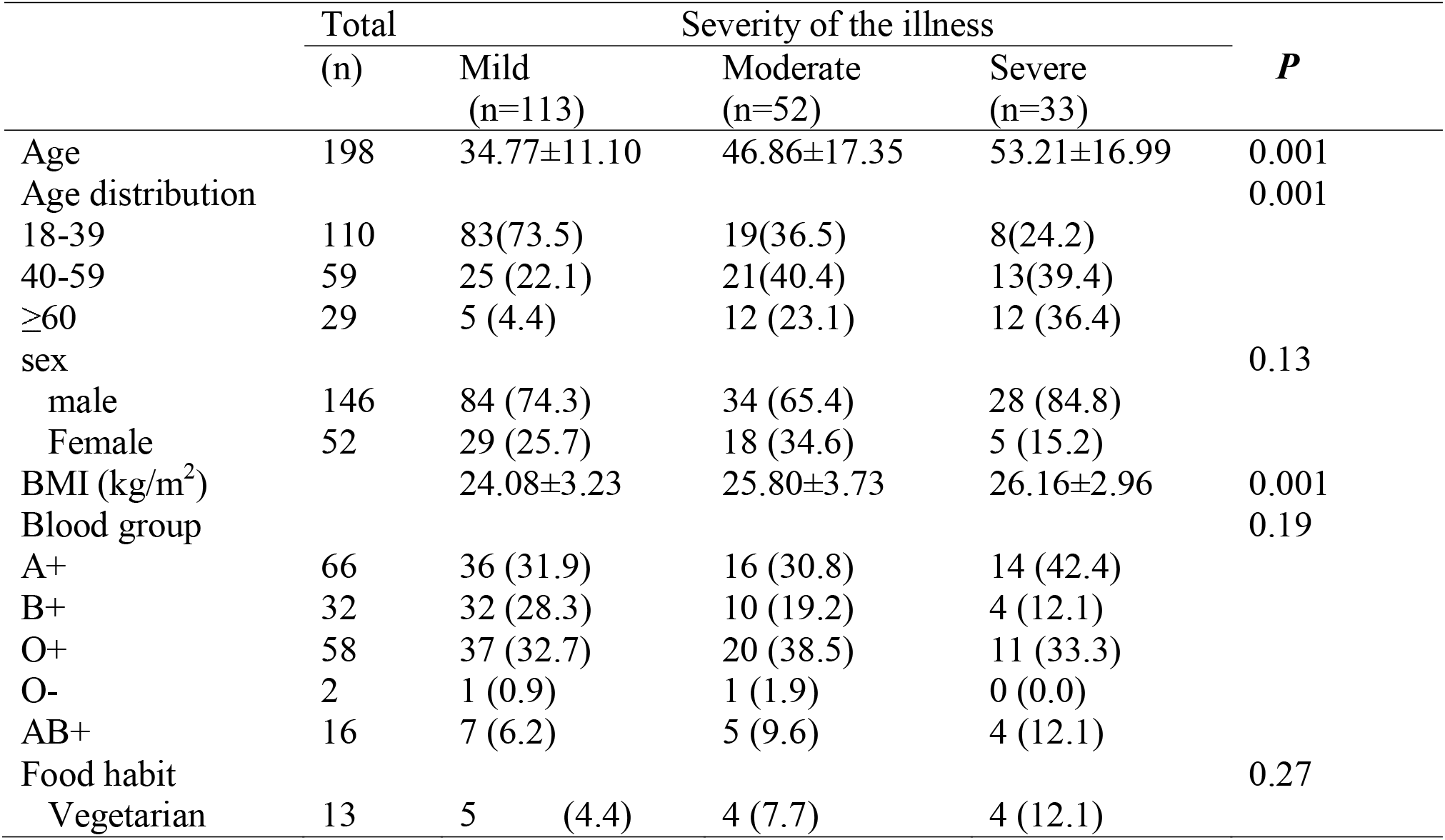
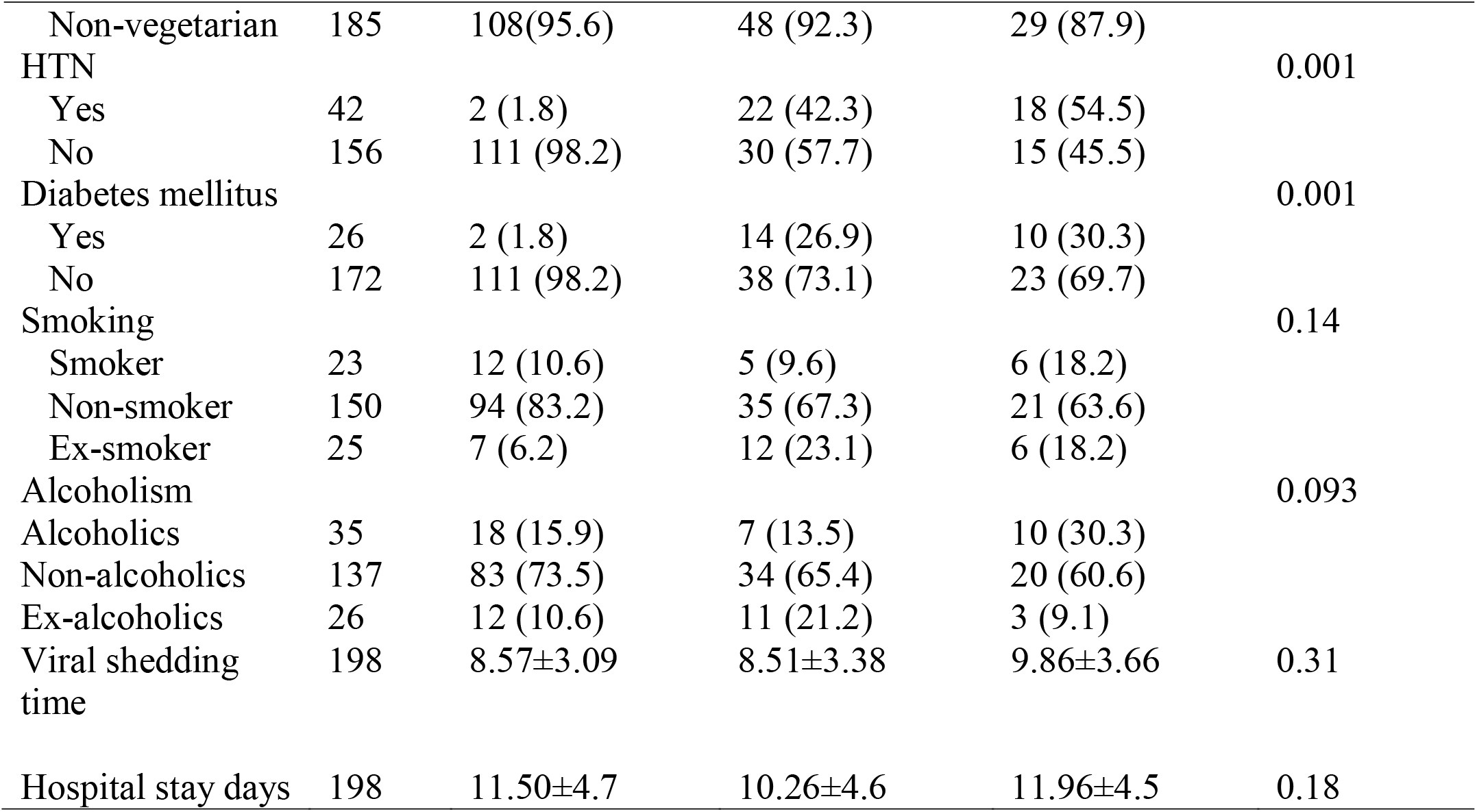
Association between severity of the disease and demographic/clinical factors

The univariate and multivariate regression model was constructed including the variables age, sex, BMI, food habit, smoking history, alcohol consumption history, hypertension, and diabetes mellitus (table 5). In univariate analysis, each 1-year increase in age (OR: 1.05; P<0.001; 95%CI:1.030-1.081), each 1 unit increase in BMI (OR:1.12; 95%CI:1.02-1.25; P:0.033), hypertension (OR:5.95; 95% CI: 2.66-13.30: P<0.001), diabetes mellitus (OR:3.26; 95%CI:1.30-8.15: P<0.005), comorbid illness (OR: 5.79; 95%CI: 2.51-13.33; P<0.001), and fever (OR:34.64; 95%CI:7.98-150.31; P<0.001) were independently associated with the severity of the illness, and each 1 unit increase in age (OR: 1.049; 95% CI: 1.019-1.080; P=0.02), hypertension (OR: 4.77; 95%CI: 1.62-14.04; P=0.004), and fever (OR: 51.02; 95%CI: 9.56-272.51; P<0.001) remained a significant factor in multivariate analysis.

**Table 5.** logistic regression analysis of factors associated with the severity of illness

|  | Univariate model |  | Multivariate model |  |
| --- | --- | --- | --- | --- |
|  | OR; 95%CI | P | OR; 95%CI | P |
| Age, years | 1.056 (1.030 -1.081) | <0.001 | 1.049 (1.019-1.080) | 0.002 |
| **Sex** |  |  |  |  |
| Male | 2.23 (0.81 -6.12) | 0.11 | 3.308 (0.987-11.08) | 0.053 |
| Female | Reference | - | - | - |
| BMI (kg/m<sup>2</sup>) | 1.12 (1.01-1.25) | 0.033 | 1.06 (0.92-1.21) | 0.41 |
| **Food habit** |  |  |  |  |
| Vegetarian | 2.39 (0.69 -8.28) | 0.16 | 0.49(0.11-2.25) | 0.36 |
| Non-vegetarian | Reference | - | - | - |
| **Smoking** |  |  |  |  |
| Current smoker | 0.88 (0.22 -3.39) | 0.85 | 1.41 (0.32-6.970) | 0.61 |
| Non-smoker | 0.54 (0.19-1.51) | 0.24 | 2.56 (0.68-9.57) | 0.16 |
| Ex-smoker | Reference | - | - | - |
| **Alcoholism** |  |  |  |  |
| Alcoholics |  |  | 3.30(0.55-19.50) | 0.18 |
| Non-alcoholics |  |  | 3.68(0.72-18.73) | 0.11 |
| Ex-alcoholics | Reference |  | - | - |
| **Hypertension** |  |  |  |  |
| Present | 5.95 (2.66-13.30) | 0<0.001 | 4.77 (1.62-14.04) | 0.004 |
| Absent | Reference | - | - | - |
| **Diabetes mellitus** |  |  |  |  |
| Present | 3.26 (1.30 -8.15) | 0<0.001 | 0.76(0.23-2.48) | 0.65 |
| Absent | Reference | - | - | - |
| **Comorbidity\*** | 5.79 (2.51 -13.33) | 0<0.001 | 2.05(0.54-8.01) | 0.25 |
| Present |  |  |  |  |
| Absent |  |  |  |  |
| **Fever** |  |  |  |  |
| Present | 34.64(7.98-150.31) | 0<0.001 | 51.02(9.56-272.51) | 0<.001 |
| Absent | Reference | - | - | - |

Note: \* presence of at least one co-existing illness

## Discussion

Identifying risk factors that contribute to severe and fatal disease progression is vital for clinicians and the public health approach to deal with the disease. Additionally, early screening of such predictive factors could contribute to research into pathophysiological processes of Covid-19 from which further treatment strategy could be developed. Evidence on covid-19 and its causal agent SARS-CoV-2 is rapidly accumulating, and most of the data are from resource-rich countries. In a resource-limited country like Nepal, however, some studies on the epidemiology of the disease have been documented, data on the severity and risk factors that contribute to the severity of the illness is still lacking.

The rate of the severity of the disease significantly varies across the region. In this study, 33 (16.7%) population had a severe illness which is higher than the study demonstrated in Guangzhou (10.4%) (27) and lower than the recently published data, where the percentage of severe and critically ill patients ranged from 19.8% to 49.0% in adults (28-30). This variation of the disease severity across the regions has been attributed due to many factors. it is suggested that that SARS-CoV-2 has genomic diversity (31), and it mutates through replication and may evolve under the pressure of immune surveillance in the human body, with its virulence, infectivity, and transmission being affected (31). Therefore, the virulence of SARS-CoV-2 may vary during transmission, and certain populations in different regions may also have an identification effect on it, in turn leading to different disease degrees and influencing factors of COVID-19 in different regions. In addition, the patients’ inherit characteristics, certain demographic, clinical characteristics, and patient lifestyle related factors may contribute to these variations (32).

In our study setting, age emerged as a significant risk factor linked to the severity of the illness in both univariate and multivariate analysis, which corresponds well with the more recent available epidemiological studies, where older age was associated with the higher disease severity (28, 32-35). Importantly, our study noted that the mean ages of the individuals with severe cases had significantly higher than those presented with either mild or moderate illness (53.21±16.99 years for severe vs 34.77±11.10 for mild and 46.86±17.35 for moderate). It is crucial to note that each 1-year increase in age in our study showed about a 5% increase in disease severity.

In this study, the severity of the disease was significantly associated with the age distribution of the patients, the percentage of severe/critically ill subjects in older adults (age>60 years) were notably higher (36.4%), which is in agreement with the recently published data; in adult cohorts, the percentage of severe and critically ill cases in hospitalized patients were ranged from 19.8% to 49.0% (28, 36). This increased severity in the elder population could be explained by the fact that in elderly patients, many comorbidities such as hypertension, diabetes, and CKD were treated with ACE inhibitors and angiotensin II receptor blockers, which upregulate the ACE2 receptor, thereby potentiating the risk of SARS-CoV-2 infection and the risk of disease severity (37). Therefore, given the age as a potential risk factor responsible for the progression of the disease, individuals with older ages warrant to have early screening and careful treatment for the comorbid illness.

In the present study BMI identified to be a potential risk factor for severe illness cases (OR: 1.12 (95%CI;1.01-1.25), which is also supported by the study demonstrated by Gao et al, where each 1-unit increase in BMI was also associated with a 12% increase in the risk of severe COVID-19 (unadjusted OR 1.12, 95% CI 1.01–1.23) (38). Moreover, a recently published large cohort study of COVID-19 patients noted that obese patients were at increased risk of hospitalization (adjusted relative risk [aRR]: 2.20) and severity (aRR: 2.30) (39). The evidence that supports the virologic and physiological mechanism underpinning the relationship between obesity and COVID-19 severity is poorly understood. However, it is suggested that more severe COVID-19 in patients with obesity may be the result of underlying low-grade chronic inflammation and suppression of innate and adaptive immune response (40). In addition, mechanical dysfunction associated with the severe obesity may potentiate the severity of lower respiratory tract infection and contribute to secondary infection (41). Thus, considering the poor prognosis of the disease in the presence of obesity, our study indicates that health care professionals involved in managing COVID should be cognizant of the increased likelihood of severe COVID-19.

Hypertension is widely recognized to be associated with the severity of the disease and remained more frequent comorbid illness in severe COVID-19 patients compared to non-severe patients (42, 43). Li et al, in their study suggested that hypertension was an independent predictor for severe COVID-19 (OR:2.01; P=0.003) (43). In the present study, hypertension appeared to be predictive factor associated with the severity of the illness in both univariate and multivariate model, and the likelihood of the severity of the illness in former analysis was 5.95 (95%CI: 2.66-13.30; P<0.001). And this risk of the severity in our study remained high even after adjusting for age, sex, BMI, smoking history, alcohol consumption status, and diabetes mellitus (OR:5.22 (OR: 4.77; 95% CI:1.62-14.04). It is worth noting that uncontrolled blood pressure remained an independent factor associated with in-hospital mortality, ICU admission, and all major risk for cardiovascular deaths and becomes a confounding factor for COVID-19 deaths (44-46). Therefore, given the poor prognosis of the disease with poor control hypertension, our study emphasizes that early screening and maintaining the blood pressure in individuals with COVID-19 is vital for better clinical outcomes. Moreover, as these individuals appear to be at higher risk of serious disease progression, they should be given special protection against the infection.

Diabetes is home to several infections including COVI-19. More recent studies have indicated that diabetes mellitus is a common comorbidity in COVID-19 patients (47) and is suggested to be a potential influencing factor for severe and fatal COVID-19 cases (48). In the present study, COVID-19 patients with diabetes comorbid were strongly associated with the severity of the disease, and the proportion of severe cases (30.3%) in diabetic individuals was significantly higher than those in the mild (1.8%) or moderate cases (26.9%). Moreover, univariate analysis indicated that risk of developing severe illness in diabetic patients had 3.26 times higher (OR: 3.26; 95% CI:1.30-8.15). This finding is in agreement with more recent epidemiological studies. For instance; a meta-analysis indicated that SARS-CoV-2 patients with diabetes had a higher risk of (RR:2.96; 95%CI:2.31-3.79) of severe disease or death (49). In a cohort study of COVID-19 patients from resource-rich country demonstrated that those with diabetes had an increased rate of hospital admission (OR:2.24; 95%CI:1.84-2.73) and critical illness (OR:1.24;95%CI:1.03-1.50) (50).

The more severe clinical course observed in diabetic patients could be due to the fact that the expression of ACE2 receptor of SARS-CoV-2 is appeared to be profoundly elevated in diabetic mellitus patients in lung and other tissues (51) which upregulates chronic inflammation, endothelial cell activation, and insulin resistance, in turn aggravating the inflammatory response and causing dysfunction of alveolar-capillary barrier (52). Thus, considering the severe clinical course and poor prognosis of COVID-19 in diabetic patients, proper measures that could potentially serve to curb the contractility of the infection should be adopted, and in the infected patients, the attention should be directed to proper glycemic control.

An earlier study indicated that the fever greater than 38.5°C on admission was positively correlated with the severity and mortality of COVID□19 (53). Russel B et al in their study in cancer patients with COVID-19 demonstrated that fever correlated significantly with the increased rate severity of the disease (54) in the present study, the patients presenting with fever significantly correlated with the had high rate of disease severity in both univariate and multivariate analysis. A more recent study noted that serum IL-6 levels, the major cytokine responsible for cytokine storm, were higher in COVID-19 patients with fever compared to those without fever (54), illustrating that fever may be caused by elevated IL-6. Thus, the finding suggests that fever is an important risk factor for severity and high fever might be a potential predictor for mortality of COVID-19.

## Limitations

Limiting to a single center with a relatively small sample size (n=298), this study may not be representative of other settings. However, the proportion of disease spectrum demonstrated in the study was comparable to those seen in other settings. Therefore, the sample may be reflective to patients with COVID-19 throughout the disease spectrum.

Additionally, unlike the resource–rich countries, the health care system in our setting is not well developed. Therefore, considering the economic condition of the patients, not all blood chemistry studies and routine tests were investigated, and hence, we were unable to evaluate the laboratory profile and their potential connection with the disease severity.

Moreover, socio-economic factors, lifestyle, and availability of quality medical care may have contributed to the severity of the COVID-19, which need to be addressed in a further large-scale study.

## Conclusion

A considerable number of the patients with COVID-19 had mild illness, whereas 16.7% had severe illness. In univariate model, age, BMI, hypertension, diabetes mellitus, comorbidity and temperature emerged as significant factors for the severity of illness, while age, hypertension and fever still remained significant predictor in multivariate analysis. Therefore, our study emphasize that health care professionals involved in managing COVID-19 should be aware of these culprits that increase the likelihood of the severity of the illness. A special protection to the infection and strategy to proactive interventions of at an early stage of the infection of these susceptible group is indispensible to diminish severity of the illness and enhance the clinical outcome.

## Supporting information

ethical approval letter

STROBE-Checklist

## Data Availability

Data are made available upon reasonable request

## Acknowledgement

We thank Nepal armed police force (APF) hospital, Balambu, Kathmandu, Nepal for allowing us for the study. We thank Nepal Health Research Council (NHRC) for the ethical approval of the study. We are grateful to individuals whose data were evaluated for the study. We highly appreciate all the healthcare personnel and staff for their invaluable support for this study.

## References

1. Huang C, Wang Y, Li X, Ren L, Zhao J, Hu Y, et al. Clinical features of patients infected with 2019 novel coronavirus in Wuhan, China. The lancet. 2020;395(10223):497–506.

2. Zhu N, Zhang D, Wang W, Li X, Yang B, Song J, et al. A novel coronavirus from patients with pneumonia in China, 2019. New England journal of medicine. 2020.

3. Li Q, Guan X, Wu P, Wang X, Zhou L, Tong Y, et al. Early transmission dynamics in Wuhan, China, of novel coronavirus–infected pneumonia. New England Journal of Medicine. 2020.

4. Guan W-j, Ni Z-y, Hu Y, Liang W-h, Ou C-q, He J-x, et al. Clinical characteristics of coronavirus disease 2019 in China. New England journal of medicine. 2020;382(18):1708–20.

5. Yang W, Cao Q, Qin L, Wang X, Cheng Z, Pan A, et al. Clinical characteristics and imaging manifestations of the 2019 novel coronavirus disease (COVID-19): a multi-center study in Wenzhou city, Zhejiang, China. Journal of Infection. 2020;80(4):388–93.

6. Tian S, Hu N, Lou J, Chen K, Kang X, Xiang Z, et al. Characteristics of COVID-19 infection in Beijing. Journal of infection. 2020;80(4):401–6.

7. Wu Z, McGoogan JM. Characteristics of and important lessons from the coronavirus disease 2019 (COVID-19) outbreak in China: summary of a report of 72 314 cases from the Chinese Center for Disease Control and Prevention. Jama. 2020;323(13):1239–42.

8. Chan JF-W, Yuan S, Kok K-H, To KK-W, Chu H, Yang J, et al. A familial cluster of pneumonia associated with the 2019 novel coronavirus indicating person-to-person transmission: a study of a family cluster. The lancet. 2020;395(10223):514–23.

9. Dong E, Du H, Gardner L. An interactive web-based dashboard to track COVID-19 in real time. The Lancet infectious diseases. 2020;20(5):533–4.

10. Tian S, Hu N, Lou J, Chen K, Kang X, Xiang Z, et al. Characteristics of COVID-19 infection in Beijing. Journal of Infection. 2020.

11. Wu Z, McGoogan JM. Characteristics of and important lessons from the coronavirus disease 2019 (COVID-19) outbreak in China: summary of a report of 72 314 cases from the Chinese Center for Disease Control and Prevention. Jama. 2020.

12. Onder G, Rezza G, Brusaferro S. Case-fatality rate and characteristics of patients dying in relation to COVID-19 in Italy. Jama. 2020;323(18):1775–6.

13. Giangreco G. Case fatality rate analysis of Italian COVIDLJ19 outbreak. Journal of medical virology. 2020;92(7):919–23.

14. Organization WH. Coronavirus disease (COVID-2019) situation reports. Available on: https://www.WHOInt/docs/default-source/coronaviruse/situation-reports/20200221-sitrep-32-covid-19. 2020.

15. ministry of Health and Population. https://covid19.mohp.gov.np/.

16. He Y. Translation: Diagnosis and treatment protocol for novel coronavirus pneumonia (trial version 7): National Health Commission, National Administration of Traditional Chinese Medicine. Infectious Microbes & Diseases. 2020.

17. World Health Organization. Clinical management of severe acute respiratory infection (SARI) when COVID-19 disease is suspected: interim guidance, 13 March 2020. 2020 [cited 2020 April 23]. Available from: https://apps.who.int/iris/handle/10665/331446

18. Chen N, Zhou M, Dong X, Qu J, Gong F, Han Y, et al. Epidemiological and clinical characteristics of 99 cases of 2019 novel coronavirus pneumonia in Wuhan, China: a descriptive study. The lancet. 2020;395(10223):507–13.

19. Eastin C, Eastin T. Clinical Characteristics of Coronavirus Disease 2019 in China: Guan W, Ni Z, Hu Y, et al. N Engl J Med. 2020 Feb 28 [Online ahead of print. The Journal of Emergency Medicine. 2020;58(4):711.

20. Kraemer MU, Yang C-H, Gutierrez B, Wu C-H, Klein B, Pigott DM, et al. The effect of human mobility and control measures on the COVID-19 epidemic in China. Science. 2020;368(6490):493–7.

21. Wang D, Hu B, Hu C, Zhu F, Liu X, Zhang J, et al. Clinical characteristics of 138 hospitalized patients with 2019 novel coronavirus–infected pneumonia in Wuhan, China. Jama. 2020;323(11):1061–9.

22. Ruan Q, Yang K, Wang W, Jiang L, Song J. Clinical predictors of mortality due to COVID-19 based on an analysis of data of 150 patients from Wuhan, China. Intensive care medicine. 2020;46(5):846–8.

23. Wu C, Chen X, Cai Y, Zhou X, Xu S, Huang H, et al. Risk factors associated with acute respiratory distress syndrome and death in patients with coronavirus disease 2019 pneumonia in Wuhan, China. JAMA internal medicine. 2020;180(7):934–43.

24. Zhou F, Yu T, Du R, Fan G, Liu Y, Liu Z, et al. Clinical course and risk factors for mortality of adult inpatients with COVID-19 in Wuhan, China: a retrospective cohort study. The lancet. 2020;395(10229):1054–62.

25. World Health Organization. Clinical management of severe acute respiratory infection (SARI) when COVID-19 disease is suspected: interim guidance, 13 March 2020. 2020 [cited 2020 April 23]. Available from: https://apps.who.int/iris/handle/10665/331446.

26. National Health Commission & State Administration of Traditional Chinese Medicine. Diagnosis and treatment protocol for novel coronavirus pneumonia (trial version 7). 3 March 2020. Accessed 31 March 2020. Available from.

27. He F, Luo Q, Lei M, Fan L, Shao X, Huang G, et al. Risk factors for severe cases of COVID-19: a retrospective cohort study. Aging (Albany NY). 2020;12(15):15730.

28. Zhang Jj, Cao Yy, Tan G, Dong X, Wang Bc, Lin J, et al. Clinical, radiological, and laboratory characteristics and risk factors for severity and mortality of 289 hospitalized COVIDLJ19 patients. Allergy. 2020.

29. Zhang K, Liu X, Shen J, Li Z, Sang Y, Wu X, et al. Clinically applicable AI system for accurate diagnosis, quantitative measurements, and prognosis of covid-19 pneumonia using computed tomography. Cell. 2020.

30. Ye Z, Zhang Y, Wang Y, Huang Z, Song B. Chest CT manifestations of new coronavirus disease 2019 (COVID-19): a pictorial review. European radiology. 2020:1–9.

31. Zijie S, Yan X, Lu K, Wentai M, Leisheng S, Li Z, et al. Genomic diversity of SARS-CoV-2 in Coronavirus Disease 2019 patients. https://academicoupcom/cid/advance-article-pdf/doi/101093/cid/ciaa203/32866932/ciaa203pdf. 2020.

32. Wolff D, Nee S, Hickey NS, Marschollek M. Risk factors for Covid-19 severity and fatality: a structured literature review. Infection. 2020:1–14.

33. Zhang J, Wang X, Jia X, Li J, Hu K, Chen G, et al. Risk factors for disease severity, unimprovement, and mortality of COVID-19 patients in Wuhan, China. Clinical Microbiology and Infection. 2020.

34. Ebinger JE, Achamallah N, Ji H, Claggett BL, Sun N, Botting P, et al. Pre-Existing Characteristics Associated with Covid-19 Illness Severity. medRxiv. 2020.

35. Zhou F, Yu T, Du R, Fan G, Liu Y, Liu Z, et al. Clinical course and risk factors for mortality of adult inpatients with COVID-19 in Wuhan, China: a retrospective cohort study. The lancet. 2020.

36. Zhang K, Liu X, Shen J, Li Z, Sang Y, Wu X, et al. Clinically applicable AI system for accurate diagnosis, quantitative measurements, and prognosis of COVID-19 pneumonia using computed tomography. Cell. 2020;181(6):1423-33. e11.

37. Shahid Z, Kalayanamitra R, McClafferty B, Kepko D, Ramgobin D, Patel R, et al. COVIDLJ19 and older adults: what we know. Journal of the American Geriatrics Society. 2020;68(5):926–9.

38. Gao F, Zheng KI, Wang X-B, Sun Q-F, Pan K-H, Wang T-Y, et al. Obesity is a risk factor for greater COVID-19 severity. Diabetes care. 2020;43(7):e72–e4.

39. Fresán U, Guevara M, Elía F, Albéniz E, Burgui C, Castilla J, et al. Independent Role of Severe Obesity as a Risk Factor for COVIDLJ19 Hospitalization: A Spanish PopulationLJBased Cohort Study. Obesity. 2021;29(1):29–37.

40. Saltiel AR, Olefsky JM. Inflammatory mechanisms linking obesity and metabolic disease. The Journal of clinical investigation. 2017;127(1):1–4.

41. Dixon AE, Peters U. The effect of obesity on lung function. Expert review of respiratory medicine. 2018;12(9):755–67.

42. Li R, Tian J, Yang F, Lv L, Yu J, Sun G, et al. Clinical characteristics of 225 patients with COVID-19 in a tertiary Hospital near Wuhan, China. Journal of Clinical Virology. 2020;127:104363.

43. Li X, Xu S, Yu M, Wang K, Tao Y, Zhou Y, et al. Risk factors for severity and mortality in adult COVID-19 inpatients in Wuhan. Journal of Allergy and Clinical Immunology. 2020.

44. Ran J, Song Y, Zhuang Z, Han L, Zhao S, Cao P, et al. Blood pressure control and adverse outcomes of COVID-19 infection in patients with concomitant hypertension in Wuhan, China. Hypertension Research. 2020;43(11):1267–76.

45. Gao C, Cai Y, Zhang K, Zhou L, Zhang Y, Zhang X, et al. Association of hypertension and antihypertensive treatment with COVID-19 mortality: a retrospective observational study. European Heart Journal. 2020;41(22):2058–66.

46. Zhou D, Xi B, Zhao M, Wang L, Veeranki SP. Uncontrolled hypertension increases risk of all-cause and cardiovascular disease mortality in US adults: the NHANES III Linked Mortality Study. Scientific reports. 2018;8(1):1–7.

47. Ou M, Zhu J, Ji P, Li H, Zhong Z, Li B, et al. Risk factors of severe cases with COVID-19: a meta-analysis. Epidemiology & Infection. 2020;148.

48. Du H, Dong X, Zhang Jj, Cao Yy, Akdis M, Huang Pq, et al. Clinical characteristics of 182 pediatric COVIDLJ19 patients with different severities and allergic status. Allergy. 2020.

49. Guo L, Shi Z, Zhang Y, Wang C, Moreira NCDV, Zuo H, et al. Comorbid diabetes and the risk of disease severity or death among 8807 COVID-19 patients in China: A meta-analysis. diabetes research and clinical practice. 2020;166:108346.

50. Petrilli CM, Jones SA, Yang J, Rajagopalan H, O’Donnell L, Chernyak Y, et al. Factors associated with hospital admission and critical illness among 5279 people with coronavirus disease 2019 in New York City: prospective cohort study. Bmj. 2020;369.

51. Xinhua. Last Updated: 2020-02-14 00:36. U.S. economist says impact of COVID-19 on Chinese, world economy limited.

52. Hayden MR. Endothelial activation and dysfunction in metabolic syndrome, type 2 diabetes and coronavirus disease 2019. Journal of International Medical Research. 2020;48(7):0300060520939746.

53. WHO. Coronavirus disease (COVID-2019) situation reports. 2020.

54. Del Valle DM, Kim-Schulze S, Huang H-H, Beckmann ND, Nirenberg S, Wang B, et al. An inflammatory cytokine signature predicts COVID-19 severity and survival. Nature medicine. 2020;26(10):1636–43.

