## Supplementary material for "Predisposing factors associated with the severity of the illness in adults with Covid-19 in Nepal": ethical approval letter

Government of Nepal

### Nepal Health Research Council (NHRC)

Estd. 1991

Ref. No.: 1297

**12 November 2020**

**Dr. Roshan Kumar Jha**

Principal Investigator

Nepal APF Hospital

**Ref: Approval of research proposal**

**Dear Dr. Jha,**

This is to certify that the following protocol and related documents have been reviewed and granted approval by the Expedited Review Sub-Committee for implementation.

| | | | |
|---|---|---|---|
| **ERB Protocol Registration No.** | 723/2020 P | **Sponsor Protocol No** | NA |
| **Principal Investigator/s** | Dr. Roshan Kumar Jha | **Sponsor Institution** | NA |
| **Title** | Epidemiology and Clinical spectrum and treatment outcome of COVID-19 patients in Nepal APF Hospital Balambu Kathmandu | | |
| **Protocol Version No** | NA | **Version Date** | NA |
| **Other Documents** | 1. Data collection tools<br>2. Acceptance letter from study site | **Risk Category** | Minimal risk |
| **Expedited Review** | Proposal [√]<br>Amendment [ ]<br>Re-submitted [ ]<br>**Meeting Date:** 10 November 2020 | **Duration of Approval**<br>12 November 2020 to 12 November 2021 | **Frequency of continuing review** |
| **Total budget of research** | **NRs 25,000.00** | | |
| **Ethical review processing fee** | **NRs 5,000.00** | | |
| **Investigator Responsibilities** | | | |
| • Any amendments shall be approved from the ERB before implementing them | | | |

Ref. No.: 1297

- Submit progress report every 3 months
- Submit final report after completion of protocol procedures at the study site
- Report protocol deviation / violation within 7 days
- Comply with all relevant international and NHRC guidelines
- Abide by the principles of Good Clinical Practice and ethical conduct of the research

If you have any questions, please contact the Ethical Review M & E Section at NHRC.

**Thanking you,**

**Dr. Pradip Gyanwali**
**Member-Secretary**
**(Executive Chief)**
